# Subgrouping patients with comorbidity between functional dyspepsia and irritable bowel syndrome: application of multidimensional item response theory and latent profile analysis

**DOI:** 10.1101/2023.08.03.23293577

**Authors:** Huang Zhongyu, Lyu Zipan, Liu Fengbin

**Author notes:** Corresponding author: ZY.HUANG:;.

## Abstract

**Background:** Comorbidity between different subtypes of functional gastrointestinal disorders(FGIDs) is of high prevalence in clinical practice. Heterogeneity of clinical appearances has led to difficulty in individualized diagnosis and comprehensive management of FGIDs.

**Aims:** To discover the hidden clinical patterns of patients with comorbidity between functional dyspepsia(FD) and irritable bowel syndrome(IBS).

**Methods:** In a retrospective cross-section study, a self-report questionnaire that consist of items indicating 5 different assessing domains including gastrointestinal discomforts, systemic discomforts, psychological disorders, and environmental aggravated factors was used as basic instrument for clinical assessment. With item response theory, the theoretical framework of assessment was evaluated, and latent traits of patient were quantified in the simulated computerized adaptive testing. Latent class analysis was used for uncovering the hidden patterns over the heterogenous clinical appearances. And differences among the profiles were compared referring to the spectrum of clinical appearances and the clinical diagnosis.

**Results:** With 996 patients enrolled in the study, the validity and reliability of the instrument were evaluated as adequate (Cronbach’s alpha indices =0.72, Split-half reliability =0.84). The construct validity was also evaluated to be adequate with Chi-square/df=3.45, CFI=0.92, GFI=0.96, RMSEA=0.05, TLI=0.90, RMR=0.02. The 7-profile model was evaluated to be with better fitness with Entropy=0.98, Lo-Mendell-Rubin likelihood ratio test-*p* value<0.01, Bootstrap likelihood ratio test-*p* value<0.01. And the patterns detailed the heterogeneity of clinical appearance of FGIDs patients either in general condition or discomfort in specific dimension.

**Conclusions:** With application of multidimensional variable analysis, this article summarized the hidden patterns beneath the heterogenous clinical features. And quantitative approaches helped equip clinician with individualized and comprehensive tool in the management of complex diseases such as FGIDs.

## Introduction

Characterized by persistent and recurring gastrointestinal symptoms, functional gastrointestinal disorders (FGIDs) affected people with a prevalence over 40% around the world(1, 2). Although Rome III and IV criteria are widely used for diagnosis of FGIDs in research design and clinical practice, it leads to underdiagnosis and misdiagnosis of patients without capturing the full spectrum of their symptoms(3). The heterogeneity and variability of clinical manifestations of FGIDs pose a challenge for individualized diagnosis, which in turn hinders the attainment of satisfied efficacy of treatment(4). The prevalence of overlapping among FGIDs subtypes was also reported to be high, especially that between functional dyspepsia(FD) and irritable bowel syndrome(IBS)(5–7). Similar symptoms between IBS and FD such as abdominal pain, bloating and changes in bowel habits make it difficult to make comprehensive diagnosis and differentiation of the two conditions(8). Relying on self-reported symptoms, it is difficult to make a definitive diagnosis with clinical criteria such as Rome IV. Common pathophysiological mechanism that contributed to both FD and IBS were reported involving the dysfunction of gastrointestinal motility(9, 10), visceral hypersensitivity(11), immune activation(12), abnormal brain-gut interaction(13) and psychosocial factors(14). However, the lack of objective biomarkers makes it difficult to confirm a diagnosis with certainty. Comorbidities with different subtypes of FGIDs and non-gastrointestinal disorders reminds clinician to consider a comprehensive approach while diagnosing and treating patients with FGIDs.

Aiming at shifting towards a more individualized and comprehensive mode in treating FGIDs, in recent decades, quantitative instruments and patient-centered care models were developed for assisting decision-making in clinical practice(15). Scales were developed for measuring multidimensional clinical appearances including gastrointestinal somatic symptoms, extraintestinal somatic symptoms and psychological symptoms(16, 17). From interdisciplinary perspective, data science methods were applied to discover latent patterns beneath the complex clinical appearances of diseases(18, 19). As typical strategy, principal component analysis and latent class analysis(LCA) were normally used for identifying subgroups of FGIDs via clustering symptoms and extracting relationship among the complex clinical appearance(20–22). Advanced applications were also developed over the quantitative approaches. Informatic platforms such as Rome IV multidimensional clinical profile (MDCP) and Interactive Clinical Decision Toolkit were also developed to assist clinical decision and individualized comprehensive management of FGIDs(23–25). Although the application of these tools represent a significant paradigm shift in the field FGIDs assessment and treatment the adaptation of the assessing procedure and interpretability of the assessing results were to be optimized for further application in clinical practice(26). Moreover, due to the regional differences derived from different countries and ethnicity, it is crucial to develop patient-centered and condition adaptive instruments to meet the need of regional practice in assessing FGIDs.

Innovated testing theory and algorithm such as multidimensional item response theory (MIRT) and structural equation modelling (SEM) were applied for enhancing feasibility and interpretability of the assessment in specific occasion(27, 28).In these paradigm, conceptual framework could be constructed and validated with SEM and psychometric properties of the instrument could be estimated with MIRT. With a flexible framework, computer adaptive testing(CAT) on basis of MIRT also revealed adaptively quantification of examinees’ latent traits in an more efficient way(29, 30). Taking advantage of these multidisciplinary approaches, this article attempted to estimate an innovative methodological paradigm for individualized assessment of patients with comorbidity of FD and IBS.

## Materials and methods

### Self-report questionnaire and assessing framework

Through a comprehensive review on the clinical prevalence of FGIDs related symptoms, an item pool was constructed referring 5 regional experts’ consensus and clinical guidance about FD and IBS published in Asia(31–35). Content of items was designed referring to the setting of scales including IBS-SSS(36) and SF-36(37). Following guidance of ROME IV criteria, option setting of each item was designed and illustrated in 4-points Likert style. For the category of symptom intensity, option 0 indicates inapplicable and 1 indicates Mild degree fitting those symptoms are not quite apparent and attack once a week. For Moderate with score 2, symptoms are apparent but do not disrupt daily activities which occur 2-3 days in a week. Severe degree with score 3 implicated that symptoms are apparent and strongly interfere with daily life which occur 4-5 days or more in a week. For the non-gastrointestinal factors including dietary, environmental, and mental factors, dichotomous items were designed to discriminate inductive and alleviate factor associated with clinical discomforts.

Cultural adjustment process was carried out with a group discussion among clinical practitioners and experts. And symptoms such as “abdominal cold” and “overwork aggravated discomfort” that not specified in Rome criteria were drafted and added in the assessment considering the regional prevalence in China. Finally, the questionnaire was designed consisting of 26 items over a theoretical framework covering five latent traits including gastrointestinal symptoms, systemic symptoms, environmental sensitivity, and emotional discomforts.

### Participants

A cross-sectional study was designed for collecting clinical features of patients diagnosed with FD or IBS according to Rome IV criteria. Diagnosis was made by the senior clinic practitioner after ruling out potential organic leisons with gastrointestinal endoscopy. Patients were enrolled after consent following introduction of staff in research center from Oct 2017 to Dec 2019 in the first affiliated hospital of Chinese medicine. Severity of symptoms spectrum together with clinical diagnosis of each case were recorded with the self-administrated questionnaire for further analysis. As exclusion criteria, patients with organic disease and those with severe systemic disease or with history of drug or alcohol abusement were not enrolled in this study. Those with incapacity to cooperate throughout the research process were also eliminated ensuring the quality of response. The study protocol was approved by the Ethical Committee of the First Affiliated Hospital of Chinese Medicine (NO.AF/JD-01/05).

### Data management and Statistical Analysis

Record of the paper-based questionnaires was typed and validated in Excel with cooperation of two staffs. With validation of a third staff, conflicting records were confirmed and corrected. The tolerance of missing rate of each case was set as 5% by which cases were regarded as low-quality and to be eliminated.

Reliability of the assessment was evaluated with Cronbach’s alpha indices that calculated in SPSS 21.0. As shown in Table 1, the theoretical model was constructed consisting of 5 dimensions including systemic symptoms, upper gastrointestinal symptoms, lower gastrointestinal symptoms, environmental sensitivity and emotional disorders. The structural validity of the theoretical framework was evaluated with Confirmatory factor analysis (CFA) in AMOS 21.0. And model fitting indices were evaluated including Goodness of Fit Index(GFI), Comparative Fit Index(CFI), Root Mean Square Error of Approximation(RMSEA), Root Mean Square Residual(RMR) and Tucker Lewis Index(TLI).

**Table 1.** Framework of the theoretical model for assessment of FGIDs with 5 latent traits.

| Index | Latent traits | Symptoms or factors for assessment |
| --- | --- | --- |
| 1 | Systemic symptoms | Back Pain; Chest Pain; Joint Pain; Palpitations; Insomnia; Fatigue |
| 2 | Upper gastrointestinal symptoms | Bloating; Abdominal Cold; Heartburn; Belching; Nausea; Upper Abdominal Pain; Early Fullness |
| 3 | Lower gastrointestinal symptoms | Diarrhea; Lower Abdominal Pain; Loose Stool; Hyperactive Bowel Sounds; Urgency |
| 4 | Environmental sensitivity | Irregular Diet Trigger; Overwork Trigger; Indigestive Food Trigger; Excitant Food Trigger |
| 5 | Emotional disorders | Depression; Irritating; Anxiety; Stress |

As the basic model for the CAT development, psychometric properties of each item were estimated with MIRT in R 3.2.4. And multidimensional discrimination indices (MDISC) of each item and its standardized factor loadings on latent traits were evaluated with R package mirt(38).

On basis of the MIRT model, the CAT was developed with following setting: (i). Initialization: initial values about each dimension was set as 0 and in range of (−3,3) before the starting of the assessment. (ii)Scoring: Maximum a-posterior (MAP) algorithm was used for estimating factor score with quasi-Monte Carlo integration. (iii).Point-wise Kullback-Leibler criteria was set as the rule for item selection(39). (iv). Stopping: with a simulated procedure, the test terminated when all items were answered or SE of all latent scores were less than 0.3.

Latent profile analysis(LPA) was used for estimating subgroups of the sample based on the multidimensional latent trait scores from CAT in Mplus 7.4 and R 3.2.4. In a larger scale of assessing granularity, a latent class model on basis of original responses of items was also estimated. Gaussian Mixture modelling(40) was set as the algorithm for estimating models with different number of profiles. Goodness of fit indices were estimated and compared among models including Log-Likelihood(Loglik), Akaike Information Criteria(AIC), Bayesian Information Criteria(BIC), Sample Adjusted BIC(SABIC), Entropy, Bootstrap likelihood ratio test(BLRT) and Lo-Mendell-Rubin likelihood ratio test(LMRT). And the one with most adequate fitness indices was selected for further analysis.

Comparison between IBS and FD in demographic descriptors was evaluated using Pearson’s Chi-square analysis or ANOVA in R 3.2.4. Mann-Whitney U test was used for comparing the original response of each item between FD and IBS. Moreover, intensity of symptoms and estimated scores of latent traits in the LPA models were compared using Kruskal-Wallis H test followed up by post-hoc test for pairwise comparisons of subgroups with Nemenyi’s test.

## Result

### 1. Participants

There were 1124 patients participating in the research. And 996 cases were finally enrolled meeting the inclusion criteria with the other 128 cases eliminated out of data incompleteness. As shown in Table 2, the enrolled sample consisted of 487 IBS patients and 509 FD patients according to their primary clinical diagnosis. And female took up a larger proportion than male (53.41% vs 46.59%). However, the difference of sexual was evaluated to be none statistically significant with Pearson’s Chi-squared test(p=0.935). The average age of the sample was 38.47 with range 18 to 74. And 425 of the enrolled people suffered from FGIDs related discomforts for 1 to 3 years that took up a larger proportion than others.

**Table 2.** Demographics of the sample with FD and IBS.

| Patient cohort |  |  |
| --- | --- | --- |
| Descriptives | Number of participants | 996 |
|  | Number of female/male | 532/464 |
|  | Percentage of females | 53.41% |
|  | Age | 38.47(range 18-74) |
| Diagnosis | IBS | 487(48.98%) |
|  | FD | 509(51.10%) |
| Duration (since first diagnosis) | 0-1year | 347(34.84%) |
|  | 1-3years | 425(42.67%) |
|  | More than 3 years | 224(22.49%) |

Subgrouping by original diagnosis, clinical features were compared between FD and IBS and the results were shown in Table 3. There were statistical differences existing in the typical symptoms conspicuously for the diagnosis of FD and IBS including bloating, belching, heartburn, nausea, diarrhea, lower abdominal pain, loose stool, hyperactive bowel sounds, urgency, discomforts aggravated by irregular diet together with systemic symptoms such as back pain and joint pain with p<0.05 in the Mann-Whitney U test. Other symptoms and predisposing factors were evaluated to be none significantly different between the two groups of patients.

**Table 3.**
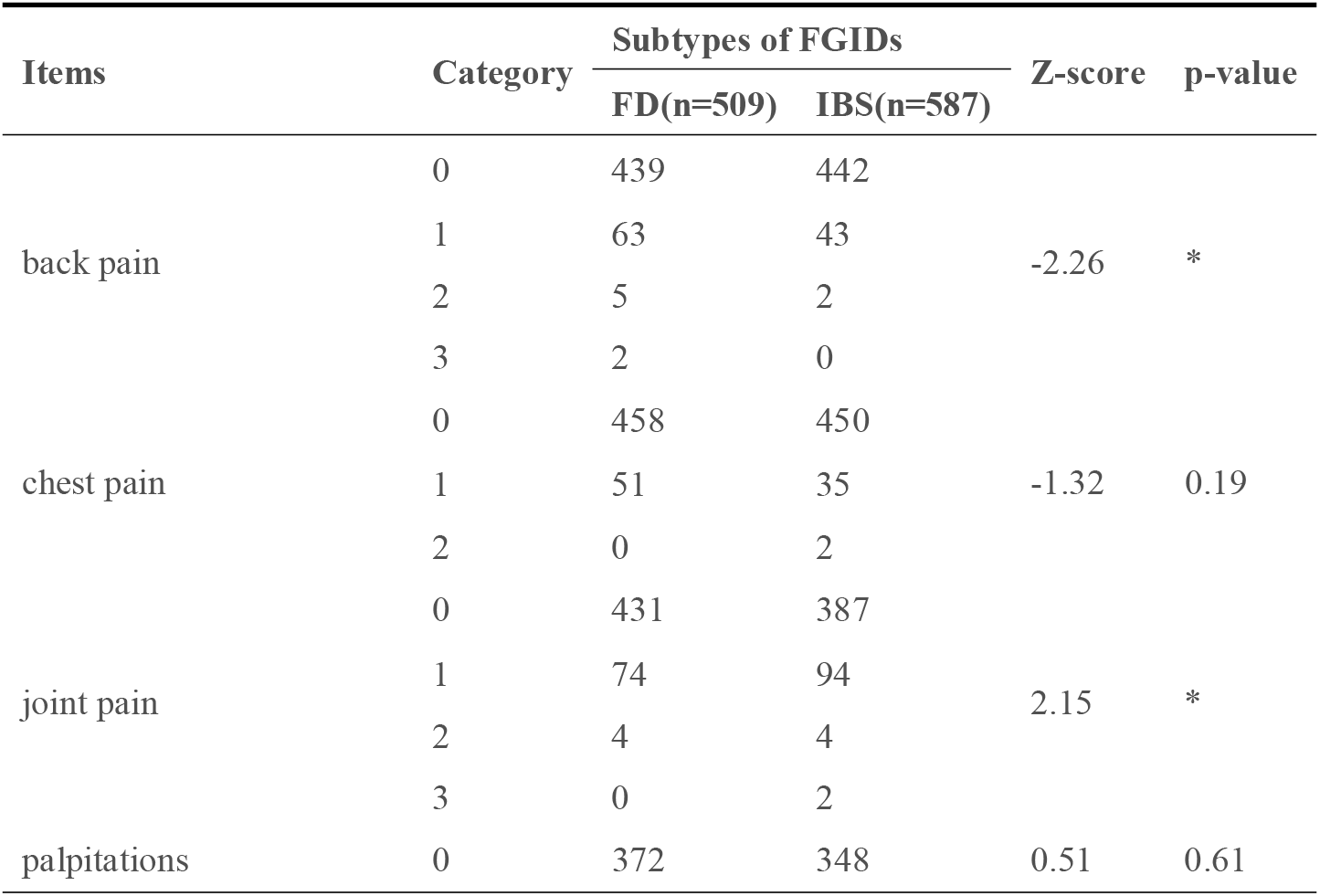
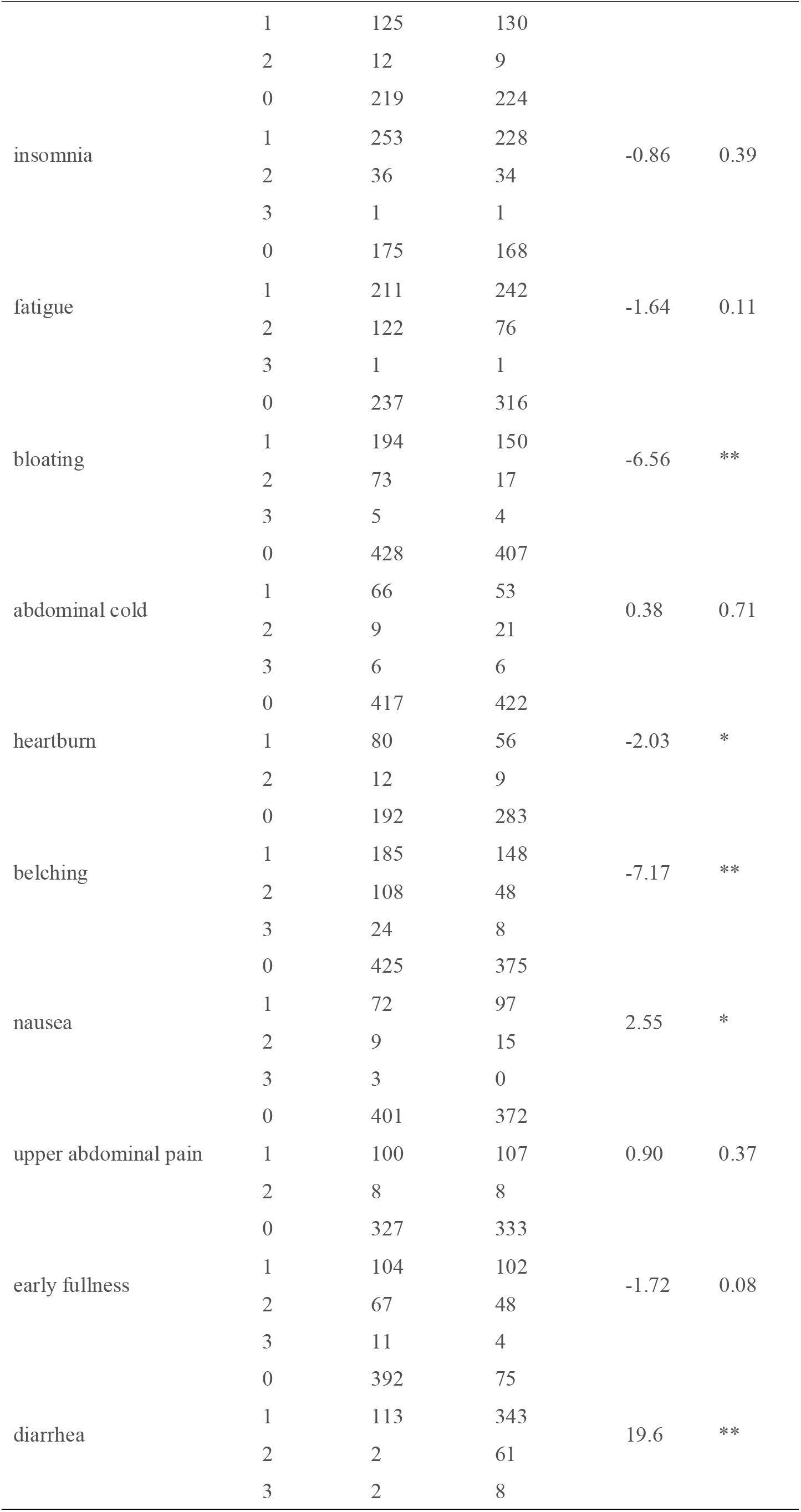
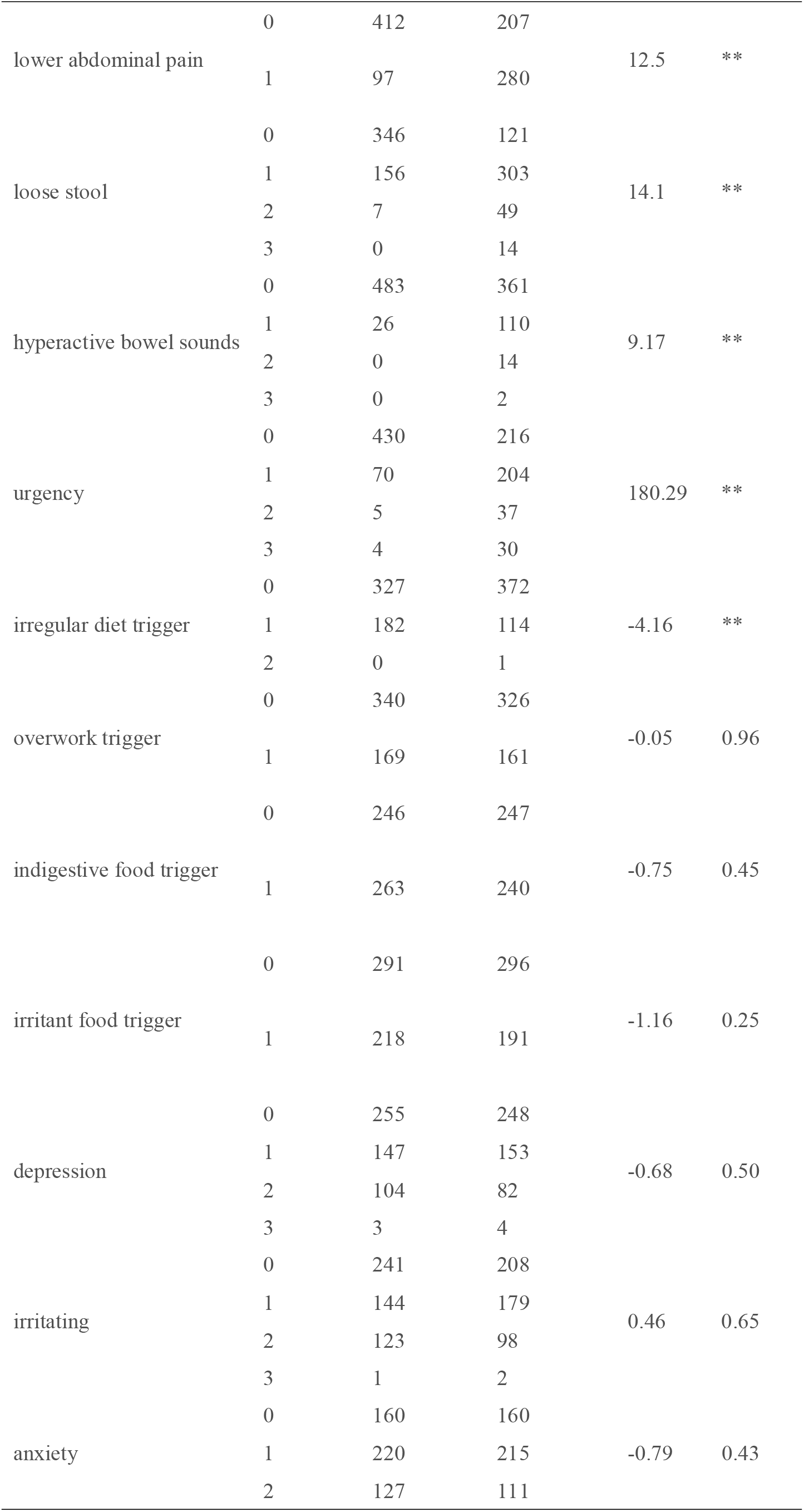
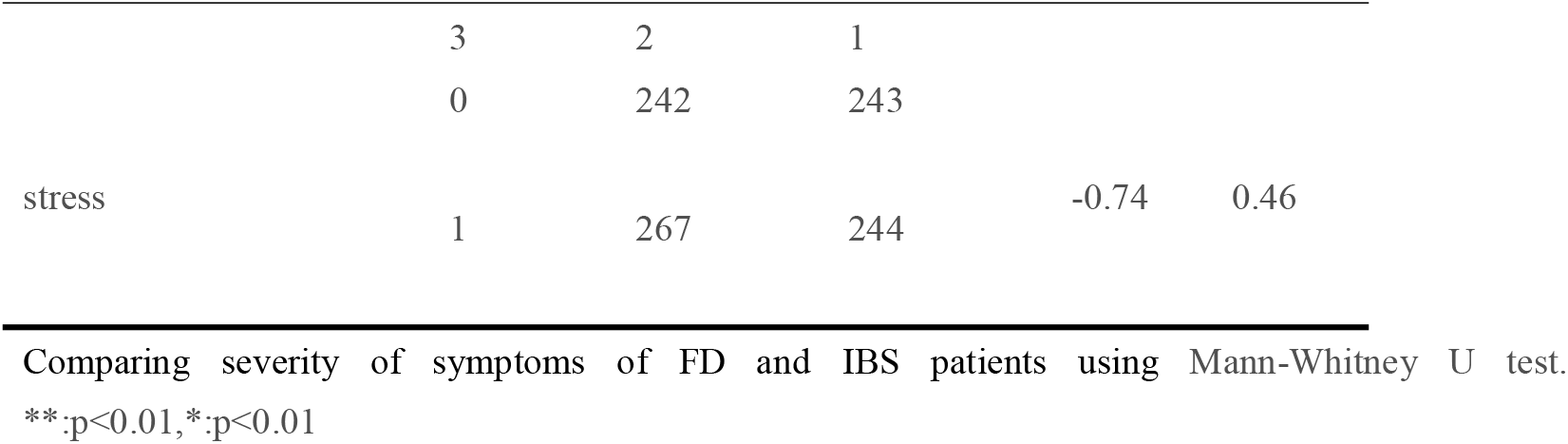
Comparison of intensity about each symptom between FD and IBS.

### 2. Validity and reliability

Validity and reliability of the self-report questionnaire was evaluated to be adequate. Cronbach’s alpha coefficient of all items of the instrument was evaluated to be 0.72 with the split half reliabilities to be 0.84. In CFA, the construct validity of the instrument was also evaluated to be adequate with Chi-square/df=3.45, CFI=0.92, GFI=0.96, RMSEA=0.05, TLI=0.90, RMR=0.02 in CFA. Further psychometric properties including MDISC and factor loadings indices of each item were evaluated in MIRT and shown in Table 4. Most items were evaluated to be with a significant loading over 0.3 with related factors. And the discrimination property of the items was evaluated to be consistent with clinical observation. For the systemic discomforts, fatigue and insomnia hold the highest factor loading indices and MDISC indices. It indicated that insomnia and fatigue could be used for characterizing patient subgroup with distinguishable property. Similarly, discrimination of bloating, nausea, abdominal cold, heartburn and early fullness as typical upper gastrointestinal discomforts were also evaluated to be adequate while belching and upper abdominal pain were in a lower level. However, comparison about three typical symptoms that known as subtype-specific between FD and IBS showed not significant differences in their severity distribution (abdominal cold(p=0.71), upper abdominal pain(p=0.37) and early fullness(p=0.08)) as shown in Table3. All lower gastrointestinal symptoms were evaluated to be with significant difference in intensity with adequate discrimination property comparing between IBS and FD. And the Z-score of symptoms in IBS were higher than that in FD subtype. As to the environmental sensitivity factor, discomforts triggered by indigestive food were evaluated with highest factor loading indicating it as the most reported discomforts of both FD and IBS. There was no difference evaluated among symptoms in psychological comorbidity factors between FD and IBS as shown in Table 3. And anxiety was evaluated with the highest factor loading while stress taking the lowest one as shown in Table4.

**Table 4.** Psychometric properties of each item of the self-report questionnaire.

| Item | MIDSC | Factor loadings |  |  |  |  |
| --- | --- | --- | --- | --- | --- | --- |
|  |  | Systemic<br>comorbidity | Upper<br>gastrointestinal<br>discomforts | Lower<br>gastrointestinal<br>symptoms | Environmental<br>sensitivity | Psychological<br>comorbidity |
| back pain | 0.47 | 0.27 | 0.00 | 0.00 | 0.00 | 0.00 |
| chest pain | 0.52 | 0.29 | 0.00 | 0.00 | 0.00 | 0.00 |
| joint pain | 0.42 | 0.24 | 0.00 | 0.00 | 0.00 | 0.00 |
| palpitations | 0.480 | 0.27 | 0.00 | 0.00 | 0.00 | 0.00 |
| insomnia | 2.96 | 0.99 | 0.00 | 0.00 | 0.00 | 0.00 |
| fatigue | 2.95 | 0.87 | 0.00 | 0.00 | 0.00 | 0.00 |
| bloating | 1.54 | 0.00 | 0.67 | 0.00 | 0.00 | 0.00 |
| abdominal cold | 1.50 | 0.00 | 0.66 | 0.00 | 0.00 | 0.00 |
| heartburn | 1.18 | 0.00 | 0.57 | 0.00 | 0.00 | 0.00 |
| belching | 0.73 | 0.00 | 0.39 | 0.00 | 0.00 | 0.00 |
| nausea | 1.66 | 0.00 | 0.70 | 0.00 | 0.00 | 0.00 |
| upper abdominal pain | 0.79 | 0.00 | 0.42 | 0.00 | 0.00 | 0.00 |
| early fullness | 1.14 | 0.00 | 0.56 | 0.00 | 0.00 | 0.00 |
| diarrhea | 4.11 | 0.00 | 0.00 | 0.92 | 0.00 | 0.00 |
| lower abdominal pain | 1.45 | 0.00 | 0.00 | 0.65 | 0.00 | 0.00 |
| loose stool | 2.55 | 0.00 | 0.00 | 0.83 | 0.00 | 0.00 |
| hyperactive bowel sounds | 2.53 | 0.00 | 0.00 | 0.83 | 0.00 | 0.00 |
| urgency | 2.31 | 0.00 | 0.00 | 0.81 | 0.00 | 0.00 |
| irregular diet trigger | 0.70 | 0.00 | 0.00 | 0.00 | 0.38 | 0.00 |
| overwork trigger | 1.01 | 0.00 | 0.00 | 0.00 | 0.51 | 0.00 |
| indigestive food trigger | 4.74 | 0.00 | 0.00 | 0.00 | 0.94 | 0.00 |
| excitant food trigger | 0.95 | 0.00 | 0.00 | 0.00 | 0.49 | 0.00 |
| depression | 2.23 | 0.00 | 0.00 | 0.00 | 0.00 | 0.79 |
| irritating | 1.94 | 0.00 | 0.00 | 0.00 | 0.00 | 0.75 |
| anxiety | 4.08 | 0.00 | 0.00 | 0.00 | 0.00 | 0.92 |
| stress | 0.65 | 0.00 | 0.00 | 0.00 | 0.00 | 0.36 |

### 3. Latent class model with original responses

To discover the hidden patterns beneath the heterogenous clinical appearances, latent class model was estimated with original intensity records of clinical symptoms. Clustering over the original response of the assessment, subgroups was defined the spectrum of symptoms in various intensity. The latent class model with 3 classes was evaluated to be adequate with p-LMRT<0.05 comparing with the 4-class model as shown in Table 5. Intensity of clinical features of each estimated subgroup were plotted in Figure 1. The first subgroup was identified with higher severity in systemic discomforts and upper gastric symptom than the other 2 subgroups. And patients in this subgroup suffered much pain and are more likely to be affected by environmental factors such as indigestive food. However, these people suffered more severe anxiety than depression. Different from Class 1, patients in the Class 2 suffered from more serious mental discomforts but less pain symptoms. Moderate level of intensity of upper gastrointestinal symptoms were evaluated as statistically significant(p<0.05) including bloating, belching, and early satisfy comparing with the other two subgroups as shown in Table 6. Upper gastric tract appearances were much typical including belching, bloating, and early fullness. Identified with higher intensity of lower gastrointestinal discomfort, patients in the Class 3 showed more serious symptoms including diarrhea, lower abdominal pain and hyperactive bowel sounds and their upper gastrointestinal symptoms were less serious than the other 2 subgroups. Higher intensity of diarrhea and lower gastrointestinal pain and lower intensity in symptoms including bloating, heartburn, belching, early fullness, and urgency were evaluated with difference(p<0.05) comparing with the Class 2.

**Figure 1.**
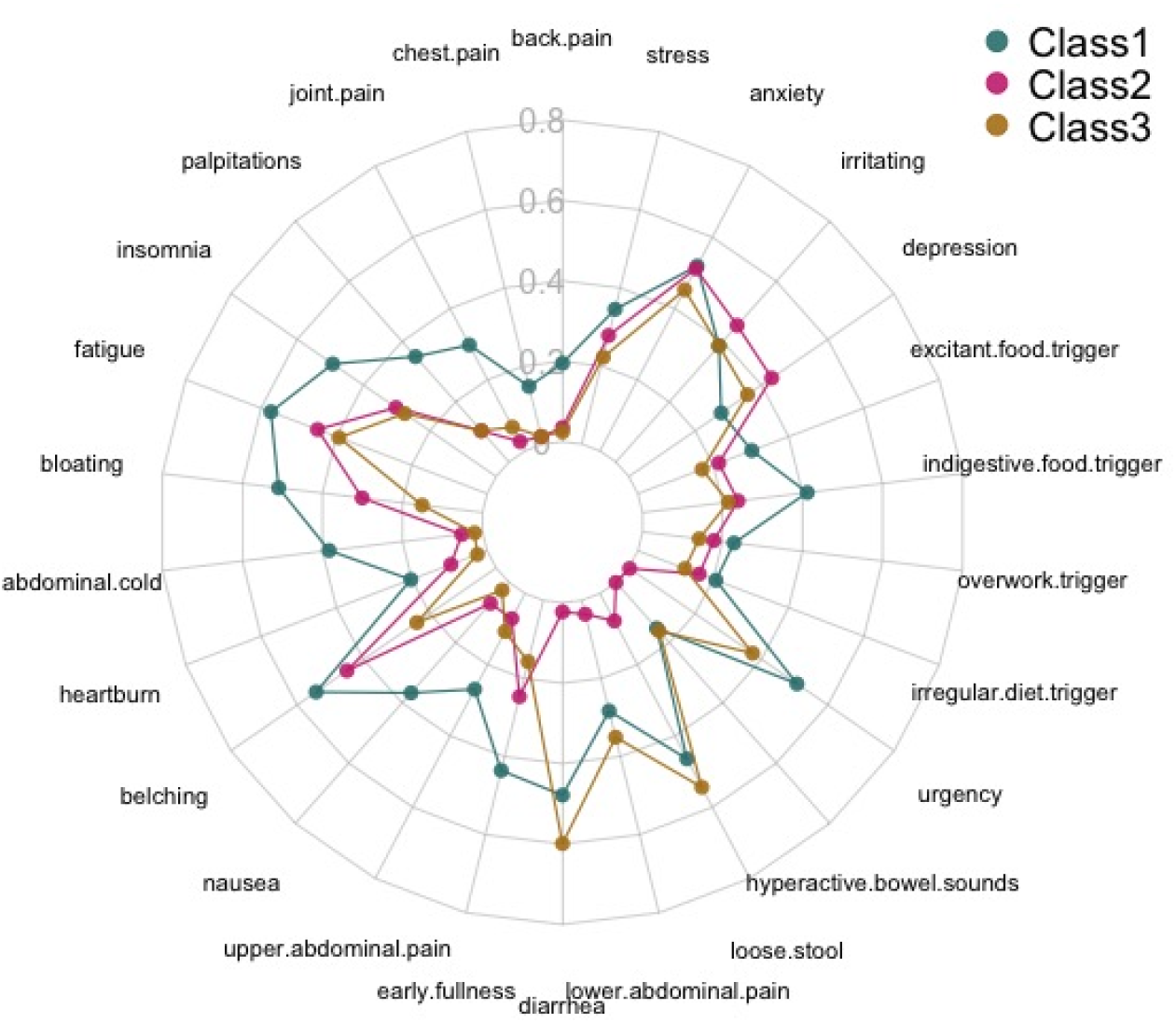
Characteristics of the 3-class latent class modelling of FD and IBS.

**Table 5.** Latent profile modelling result with computerized adaptive testing scores.

| Model | Classes | AIC | BIC | Entropy | prob_min | prob_max | n_min | n_max | BLRT_val | BLRT_p | LMRT_val | LMRT_p |
| --- | --- | --- | --- | --- | --- | --- | --- | --- | --- | --- | --- | --- |
| 1 | 1 | 38966.85 | 39275.78 | 39075.69 | 1 | 1 | 1 | 1 | - | - | - | - |
| 1 | 2 | 37374.27 | 37997.04 | 0.88 | 0.96 | 0.97 | 477/47.90% | 519/52.10% | -19420.42 | 0 | 1716.69 | <0.001 |
| 1 | 3 | 36402.54 | 37339.16 | 0.88 | 0.92 | 0.95 | 224/22.49% | 422/42.37% | -18581.24 | 0 | 1139.36 | <0.001 |
| 1 | 4 | 35894.10 | 37144.55 | 0.86 | 0.90 | 0.95 | 207/20.78% | 311/31.23% | -18019.99 | 0 | 654.41 | 0.73 |

**Table 6.**
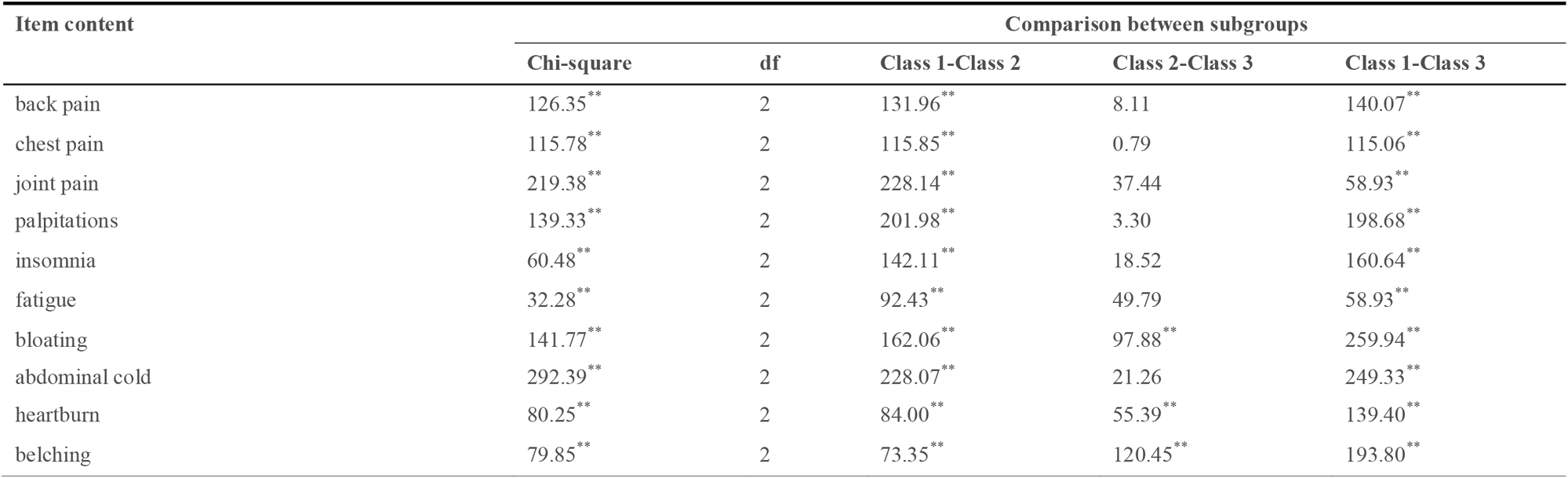
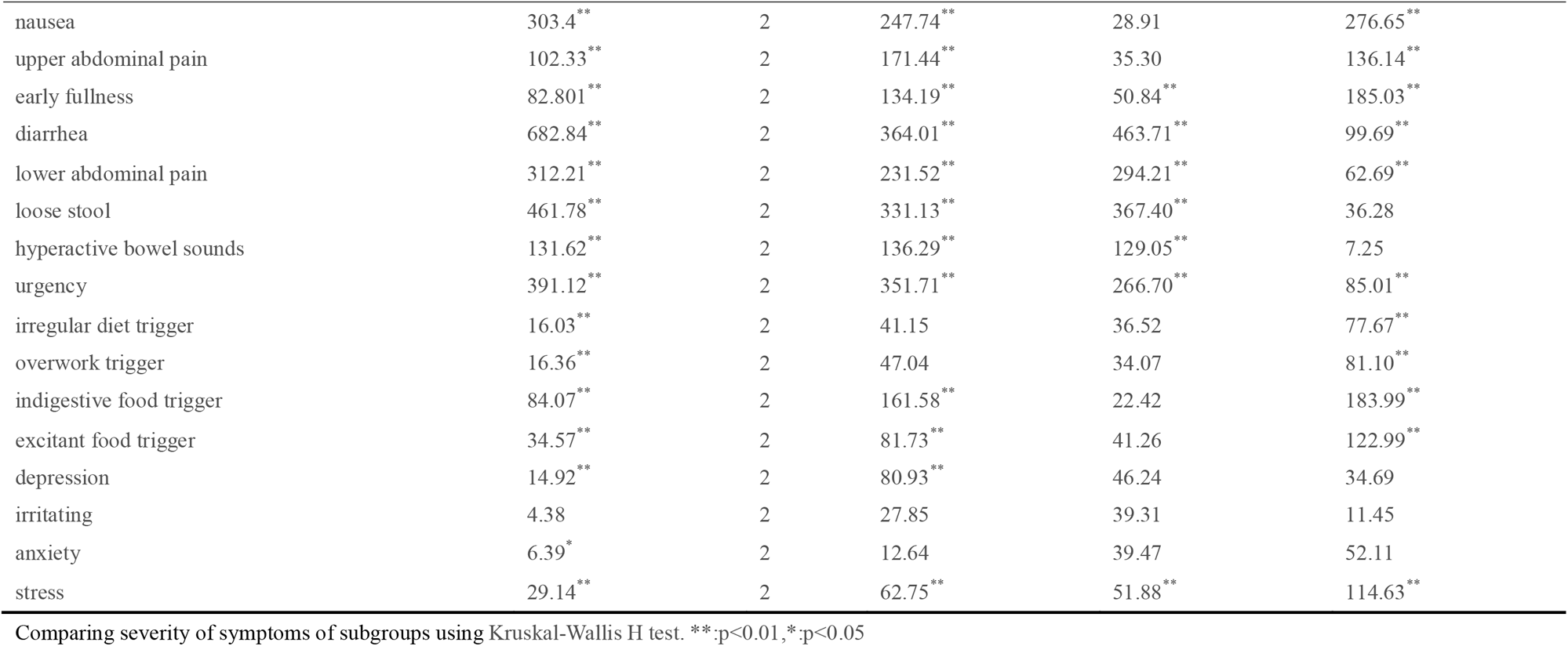
Comparison of symptom severity among the 3-class subgroups estimated with latent profile analysis.

| Item content | Comparison between subgroups |  |  |  |  |
| --- | --- | --- | --- | --- | --- |
|  | Chi-square | df | Class 1-Class 2 | Class 2-Class 3 | Class 1-Class 3 |
| back pain | 126.35** | 2 | 131.96** | 8.11 | 140.07** |
| chest pain | 115.78** | 2 | 115.85** | 0.79 | 115.06** |
| joint pain | 219.38** | 2 | 228.14** | 37.44 | 58.93** |
| palpitations | 139.33** | 2 | 201.98** | 3.30 | 198.68** |
| insomnia | 60.48** | 2 | 142.11** | 18.52 | 160.64** |
| fatigue | 32.28** | 2 | 92.43** | 49.79 | 58.93** |
| bloating | 141.77** | 2 | 162.06** | 97.88** | 259.94** |
| abdominal cold | 292.39** | 2 | 228.07** | 21.26 | 249.33** |
| heartburn | 80.25** | 2 | 84.00** | 55.39** | 139.40** |
| belching | 79.85** | 2 | 73.35** | 120.45** | 193.80** |
| nausea | 303.4<sup>**</sup> | 2 | 247.74<sup>**</sup> | 28.91 | 276.65<sup>**</sup> |
| upper abdominal pain | 102.33<sup>**</sup> | 2 | 171.44<sup>**</sup> | 35.30 | 136.14<sup>**</sup> |
| early fullness | 82.801<sup>**</sup> | 2 | 134.19<sup>**</sup> | 50.84<sup>**</sup> | 185.03<sup>**</sup> |
| diarrhea | 682.84<sup>**</sup> | 2 | 364.01<sup>**</sup> | 463.71<sup>**</sup> | 99.69<sup>**</sup> |
| lower abdominal pain | 312.21<sup>**</sup> | 2 | 231.52<sup>**</sup> | 294.21<sup>**</sup> | 62.69<sup>**</sup> |
| loose stool | 461.78<sup>**</sup> | 2 | 331.13<sup>**</sup> | 367.40<sup>**</sup> | 36.28 |
| hyperactive bowel sounds | 131.62<sup>**</sup> | 2 | 136.29<sup>**</sup> | 129.05<sup>**</sup> | 7.25 |
| urgency | 391.12<sup>**</sup> | 2 | 351.71<sup>**</sup> | 266.70<sup>**</sup> | 85.01<sup>**</sup> |
| irregular diet trigger | 16.03<sup>**</sup> | 2 | 41.15 | 36.52 | 77.67<sup>**</sup> |
| overwork trigger | 16.36<sup>**</sup> | 2 | 47.04 | 34.07 | 81.10<sup>**</sup> |
| indigestive food trigger | 84.07<sup>**</sup> | 2 | 161.58<sup>**</sup> | 22.42 | 183.99<sup>**</sup> |
| excitant food trigger | 34.57<sup>**</sup> | 2 | 81.73<sup>**</sup> | 41.26 | 122.99<sup>**</sup> |
| depression | 14.92<sup>**</sup> | 2 | 80.93<sup>**</sup> | 46.24 | 34.69 |
| irritating | 4.38 | 2 | 27.85 | 39.31 | 11.45 |
| anxiety | 6.39<sup>*</sup> | 2 | 12.64 | 39.47 | 52.11 |
| stress | 29.14<sup>**</sup> | 2 | 62.75<sup>**</sup> | 51.88<sup>**</sup> | 114.63<sup>**</sup> |

Comparing severity of symptoms of subgroups using Kruskal-Wallis H test. \*\*:p<0.01,\*:p<0.05

Clinical diagnosis of these 3 subgroups were also evaluated as the result shown in Table 7. For the Class 1, FD and IBS took up proportions with 39.73% and 60.27%. And FD took up proportion of 84.60% in the Class 2 while IBS taking up 82% in the Class 3. The proportion of diagnosis among the subgroups was evaluated to be of significant difference in the Pearson’s Chi-square analysis(p<0.001) as shown in Table 7. The pattern of clinical appearances and mixture of subtypes in the groups indicated the possible incompleteness of diagnosis of patients with more than one FGIDs subtypes. And it is also important to consider the comprehensive spectrum of symptoms so that we could better interpret the complexity of gut-brain interaction from the clinical cases.

**Table 7.** Proportion and Comparison of the clinical diagnosis of the 3-class latent class model.

| Group ID | n | Proportion | FD | IBS | Chi-square | P value |
| --- | --- | --- | --- | --- | --- | --- |
| Class 1 | 224 | 22.49% | 89 | 135 | 354.54 | <0.01 |
| Class 2 | 422 | 42.37% | 357 | 65 |  |  |
| Class 3 | 350 | 35.14% | 63 | 287 |  |  |

### 4. Latent profiles modelling with multidimensional trait scores

Summarizing different discomforts into 5 latent traits, multidimensional scores of each patient were estimated and scaled in the stimulated CAT as shown in Table 8. As all latent traits were estimated in a continuous format, LPA was used to clustering subgroups of the patients. As shown in Table 9, the model with 7 profiles was evaluated as adequate with lower BIC value. And the value of LMRT was evaluated as −197.71 (p>0.05) as comparison between the 7-profile model to the 8-profile model. Referring to the proportion of clinical subtypes, difference among the 7 subgroups was evaluated to be without statistical significance with Chi-square=11.391(*p*=0.08) as shown in Table 10. In total, there were 875 patients in subgroup P1, P2, P3 and P6 taking up 87.85% of the sample accumulatively. Different patterns of the latent traits could be observed among the 7 subgroups of FGIDs characterized by different scores of latent traits as shown in Figure 2.

**Figure 2.**
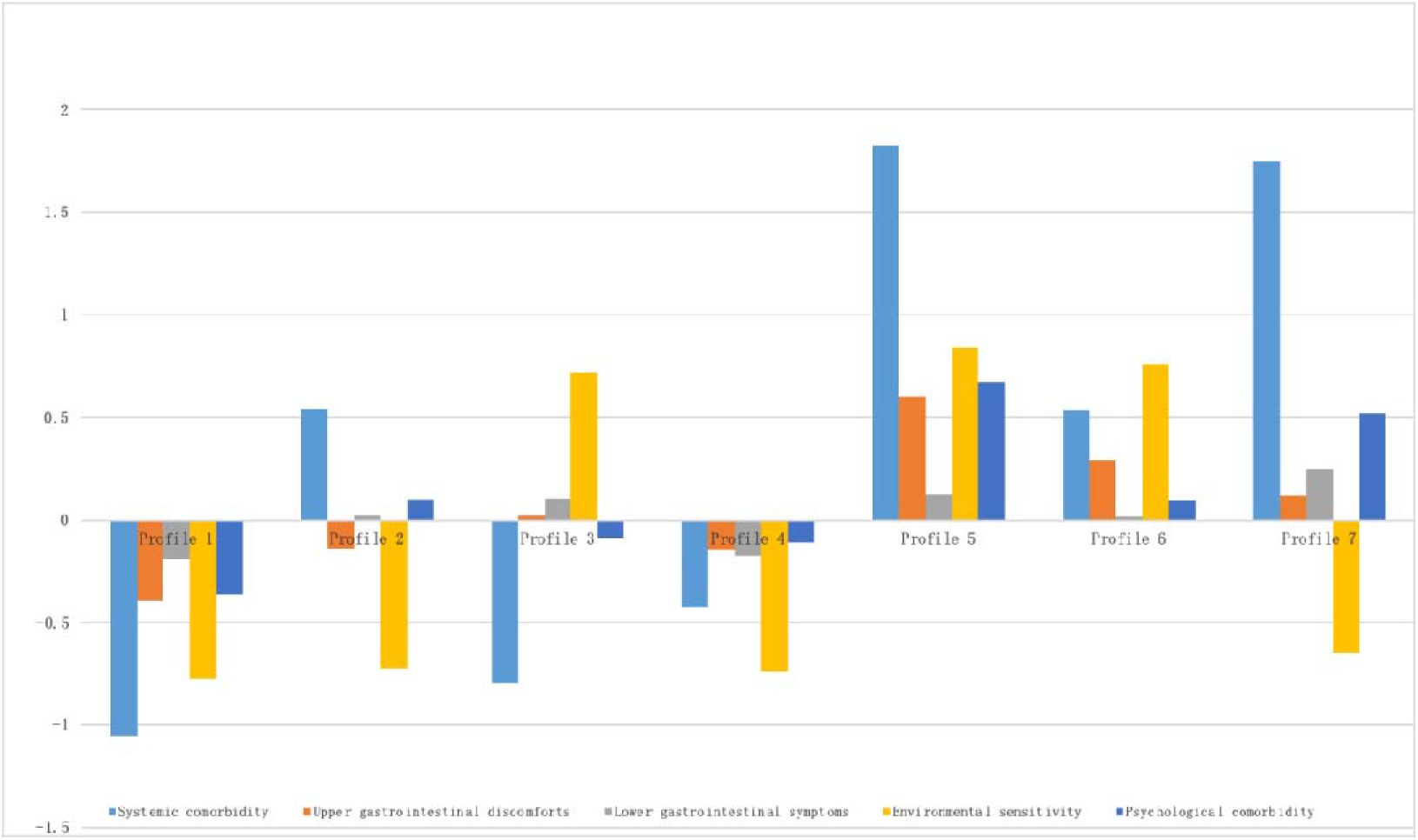
Latent traits scores of the 7-profile latent profile modelling of FD and IBS.

**Table 8.** Property of the latent traits scores estimated in the computerized adaptive testing.

| Latent trait | Minimum | Mean | Maximum |
| --- | --- | --- | --- |
| Systemic comorbidity | -1.29 | 0.00 | 2.98 |
| Upper gastrointestinal discomforts | -1.15 | 0.00 | 2.07 |
| Lower gastrointestinal symptoms | -1.19 | 0.00 | 2.65 |
| Environmental sensitivity | -1.12 | 0.01 | 1.92 |
| Psychological comorbidity | -1.38 | 0.00 | 2.99 |

**Table 9.**
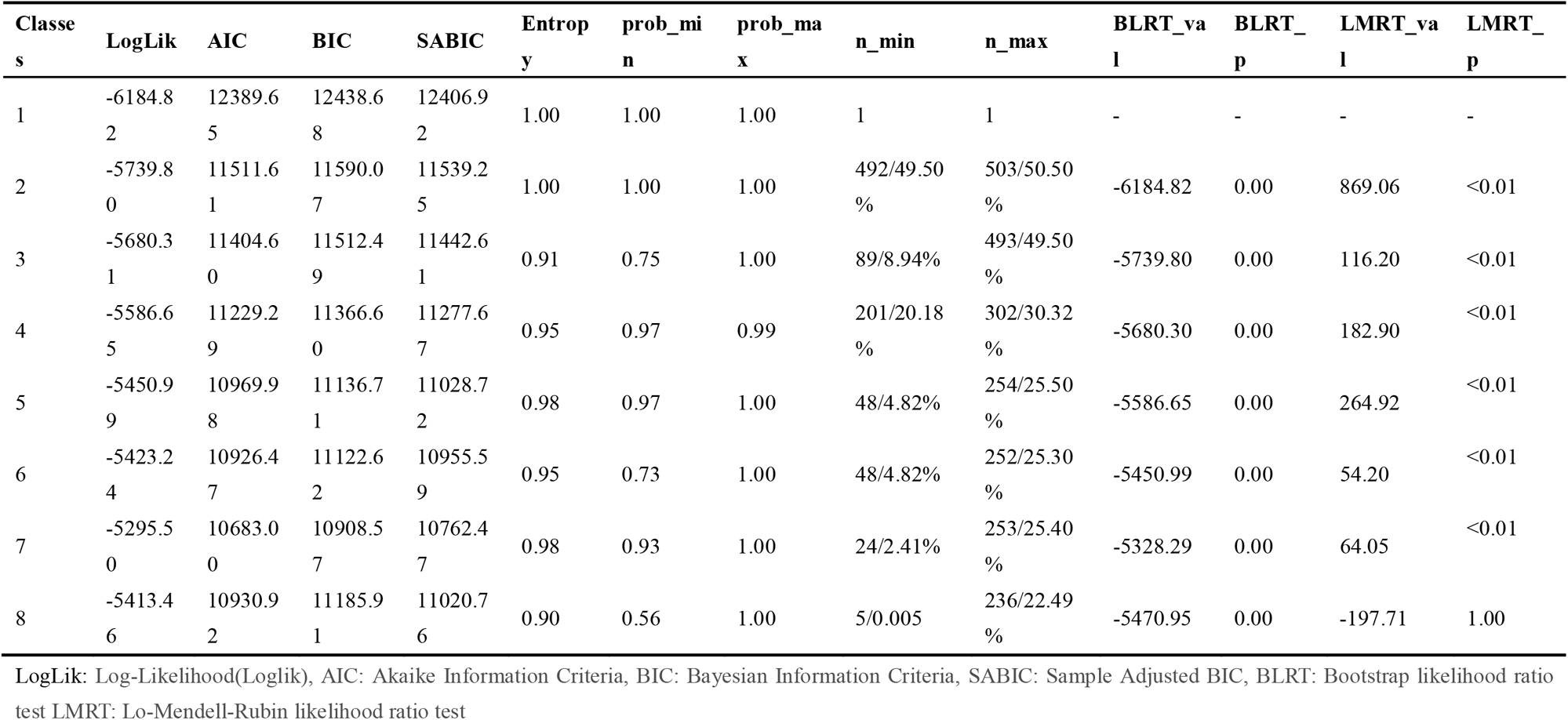
Fitness indices of the latent profile models with multidimensional computerized adaptive testing scores.

**Table 10.** Proportion and Comparison of the clinical diagnosis of the 7-profile latent profile model.

| Subgroup ID | n | Proportion | FD | IBS | Chi-square | p-value |
| --- | --- | --- | --- | --- | --- | --- |
| 1 | 192 | 19.28% | 92 | 100 |  |  |
| 2 | 228 | 22.89% | 117 | 111 |  |  |
| 3 | 202 | 20.28% | 97 | 105 |  |  |
| 4 | 49 | 4.92% | 30 | 19 | 11.391 | 0.08 |
| 5 | 48 | 4.82% | 30 | 18 |  |  |
| 6 | 253 | 25.40% | 136 | 117 |  |  |
| 7 | 24 | 2.41% | 7 | 17 |  |  |

**Table 11.**
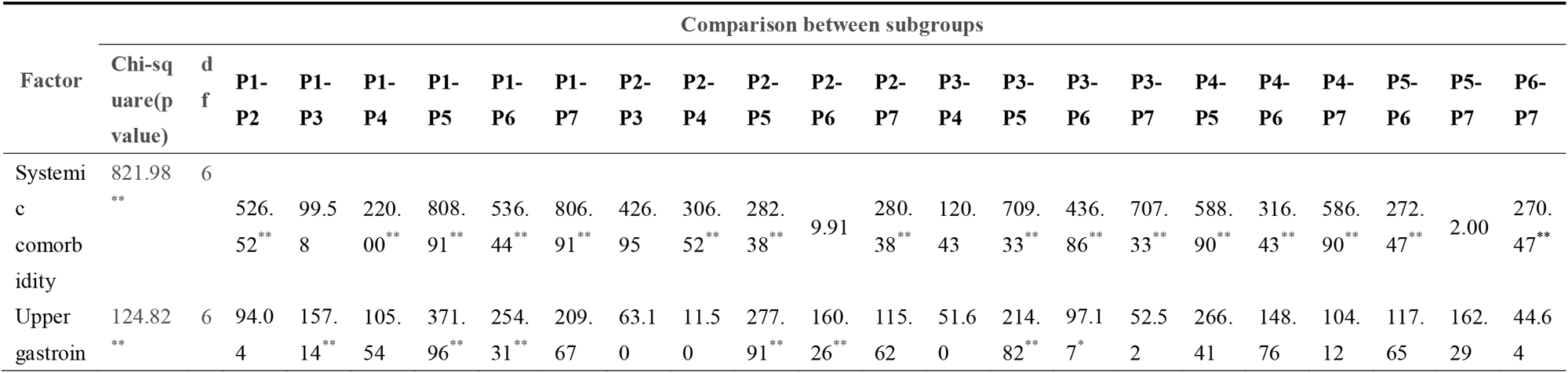
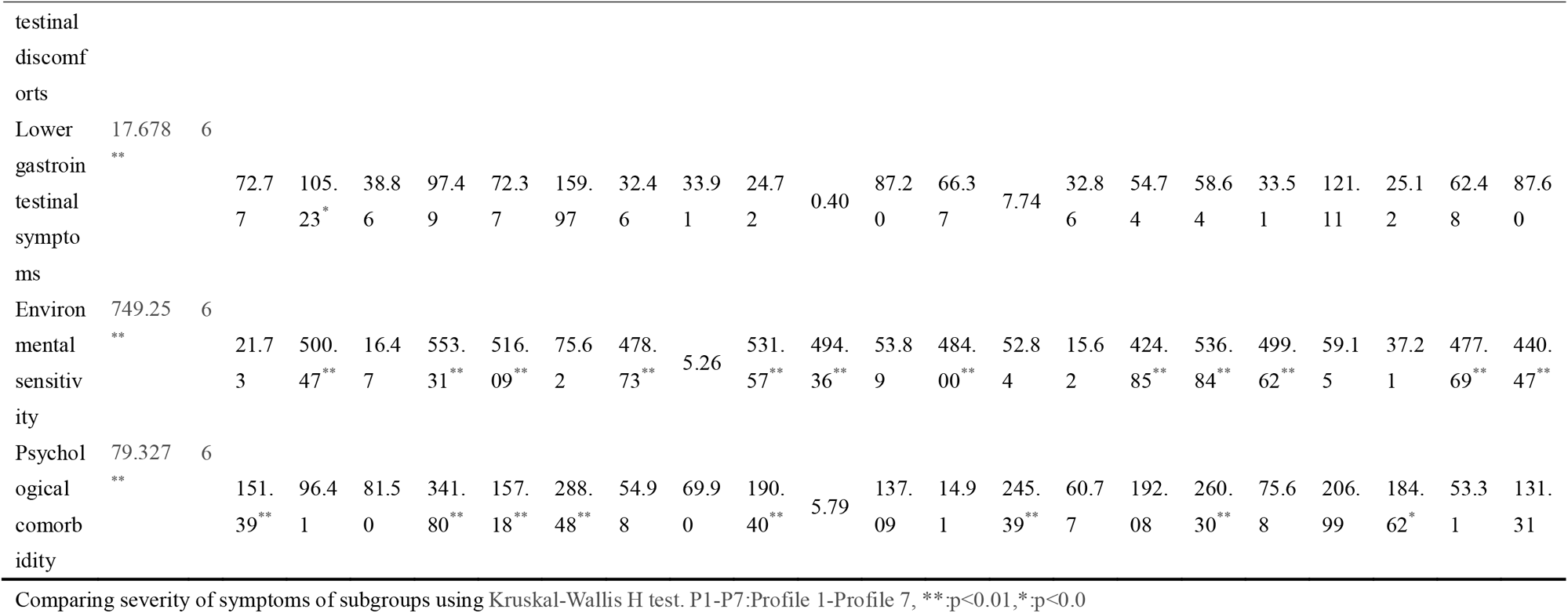
Comparison of latent traits severity among the 7-profile model with latent profile analysis.

Heterogeneity of the patterns reflected in both overall condition of the latent traits scores and intensity of distinct appearance. For example, in both the P1 and the P4 subgroups, general scores of latent traits were evaluated to be lower comparing to other subgroups. And multidimensional severity of the P5 subgroup was evaluated with a generally higher condition. As to the P2 subgroup, score of systemic symptoms was higher than those of other traits. And patients in the P3 subgroup were assessed with a high score in the environmental sensitivity dimension. In P5, P6 and P7 subgroups, higher score in GI discomforts were assessed comparing with the average condition of the subgroups. It could be observed that score of mental discomforts were positively correlated with that of the systemic discomforts in all subgroups. The differences among these 7 subgroups indicated the heterogeneity of clinical appearance of FGIDs. And the covariation between different traits provided clues for interpreting the complexity of pathogenesis of FGIDs such as gut-brain interaction.

## Discussion

Although the Rome criteria provided guidance for categorizing FGIDs with typical symptom spectrum, clinical management of FGIDs is still challenging out of the heterogeneity and variability of clinical appearances of patients. The criteria for categorizing FGIDs also showed limitation due to the mixture of the clinical manifestations between different subtypes. As the results shown in this study, patients with FD and IBS were reported to suffer from common discomforts including gastrointestinal symptoms, systemic symptoms, and environmental sensitivity in clinical practice. It’s also evaluated that patient with FD and IBS commonly suffered from mental discomforts including anxiety and irritating. The overlapping of symptoms between FD and IBS evaluated in this study showed consistency with that reported in previous clinical study(7, 41). Although the pathogenesis about the comorbidity is not well interpretated. Some biological hypotheses could help explaining the phenomenon. Disorder of gut-brain interact bidirectionally plays a crucial role in the pathophysiology of both FD and IBS(42). By using MRI or PET scanning, researchers have found that central nervous system alterations regionally existed in these two diseases(43, 44). Previous studies showed the homeostasis of mucosal cytokines and gut-homing T lymphocytes were disrupted in both FD and IBS(12, 45). It was also proposed that the comorbidity could be associated with hypersensitivity, motility abnormalities in gastrointestinal tract. The possible mechanism of this phenomenon could be about altered function of the hypothalamic-pituitary-adrenal axis and the sympathetic nervous system(46).

In this study, two models with different assessing granularity were estimated by summarizing comprehensive gastrointestinal symptoms and comorbidity with systemic symptoms and emotional disorders. As poor concordance between the data-driven clustering results and the clinical diagnosis was evaluated in both models, it reminded us of the importance of making a comprehensive assessment and diagnosis of patients with FGIDs. Following Rome criteria, patients with typical anatomical discomforts could be easily categorized and diagnosis. However, heterogeneity of the spectrum and severity of discomforts reflect the need to develop individualized assessment in a proper scale to assist assessment in clinical practice. Using multidimensional variable analysis approaches, patients with FGIDs could be varied into different subgroups according to the severity of clinical appearances(47). And the patterns of clinical appearances supported the concept that FGIDs subtypes such as functional bowel disorders should be treated as clinical continuum, other than separate disorders(48). And the systemic pathogenesis should be well inferred and summarized for better management of FGIDs other than following the anatomical classification and diagnosis criteria rigidly(49).

Heterogeneity and instability of clinical appearance also made it difficult to establish a comprehensive criteria or instrument with adequate stability and interpretability for FGIDs diagnosis. Prevalence of clinical features were reported to be different between different regions. Fluctuation of FGIDs symptoms brought difficulty for individual evaluation and treatment decision in clinical practice. As a regional specific symptom of the study, “abdominal cold” was recorded with high prevalence in China. It was also normally seen that patients without medical knowledge would report ambiguous position or property. That bring difficulty in clinical assessment and reminded us of the importance of the civilization background for the designation of instrument.

Relying on the similarity about symptoms spectrum and their severity, LPA and LCA served as data-driven methods for discovering patterns of clinical appearances of patients with comorbidity between FD and IBS. The composition of FGIDs subtype in the 3-class LCA model indicated the impracticability of Rome criteria in comprehensive assessment of patients. Neither FD nor IBS could be well described with the spectrum of the symptoms in each subgroup. The patterns of clinical appearances estimated in this study also indicated the complexity about the pathophysiology of FD and IBS. As shown in the 7-class LPA model, not only the severity of gastrointestinal symptoms but also the comorbidity with psychological and systemic discomforts should be evaluated as clues for the subgrouping patients with FD and IBS. Although the interaction between somatic discomfort and mental discomfort was not well evaluated in this study due to the lack of data from cohort study, covariation of the scores between mental discomfort and systemic discomfort could be observed. It was reported that the gut-brain interaction could serve as an important pathogenesis in the development of FGIDs. And it has become a consensus of clinical practitioners to consider the impact of mental disorders while treating patients with FGIDs. The analytic paradigm raised in this study revealed discovery of the hidden pattern of clinical appearances of patients with comorbidity between FD and IBS. However, interpretation of the model is limited due to the lack of representative data from cohort design. The model should be further optimized for better representing the pathogenesis and possible evolution among different FGIDs subgroups.

Although there were many instruments developed for subgrouping FGIDs subtypes in form of scales(18, 50), the compensatory of calculation with accumulating response scores overlaid the details about individuals and further diminished the interpretation of the assessment. With development of CAT, multidimensional latent traits were quantified as summarized mixture of symptoms. Collectively focusing on the universe of clinical appearances may yield significant benefits for FGIDs patients with comprehensive individualized instruments for clinical assessment. With the CAT estimated on the 5-dimensional MIRT model, latent traits of each patient were evaluated for upper and lower gastrointestinal discomforts, severity of emotional disorder, systemic severity and environmental sensitivity. It benefited the practitioner with comprehensive individual evaluation in a more interpretable conceptual framework. Moreover, via comparison between the 3-class model and the 7-profile model, the paradigm also provide flexible setting of granularity of assessment that could be adaptively optimized to fit the need of clinical practice.

With integrative application multivariable analysis methods, this article proposed an innovative paradigm for subgrouping FGIDs with multi-scale assessing framework. Taking personal traits other than symptoms clusters as parameters, the interpretation of the model was also strengthened for clinical practice. However, there are several limitations in our research. Firstly, a cross-section study was designed and all patients enrolled in this study were from single research center. More convincible conclusion could be drawn with a more representative sample of FGIDs patients. Further research should also be carried out for validating the stability, rationality, and further extrapolation of the model. Secondly, although the self-report questionnaire was designed via a strict procedure, the setting of items together with their psychometric properties should be optimized and re-validated in further research. In this way, the interpretability and extrapolation of the latent profile models could be modified via reducing bias that introduced from an unrepresentative sample. Thirdly, the generalizability of the findings also limited since all data was collected from China and some culturally specific items were designed such as “abdominal cold” was designed for assessment. Last but not the least, assessing logic about the CAT could also be optimized by analyzing and comparing the time cost and consistency among different settings of assessment.

## Conclusion

Making a comprehensive assessment about clinical features of FGIDs patients, this study raised a new subgrouping model for FD and IBS making use of multivariable analysis method such as CAT and LPA. The heterogenous appearances of FD and IBS patients was profiled into 7 subgroups according to the severity and the spectrum of symptoms including gastrointestinal symptoms, systemic symptoms, and environmental factors. The results suggested that criteria together with instruments need to be updated so as to make comprehensive individualized assessment of FGIDs patients. Furthermore, interdisciplinary strategy benefited clinicians with quantitative and automatic approaches for comprehensive evaluating and managing FGIDs patients which we believed would shift the clinical practice to a much more individualized mode with flexibility and availability.

## Declarations

### Ethics approval and consent to participate

All patients enrolled in this study signed informed consent. And this work was approved by the Clinical Research and Ethics Committee at the First affiliated Hospital of Guangzhou University of Chinese Medicine. (NO.AF/JD-01/05)

### Authors’ Contributions

ZY.H and ZP.L contributed toward the concept, data analysis, and manuscript writing, and manuscript review; ZY.H and ZP.L contributed toward patient and data collection; ZY.H, ZP.L, and FB.L contributed toward funding, concept, and manuscript review.

## Acknowledgments

The authors sincerely acknowledge the Guangzhou library for offering digital materials and rooms as essential support for the study.

## Data Availability Statement

The data used to support the findings of this study are available from the first author(mail) upon request.

## STROBE Statement—Checklist of items that should be included in reports of ***cross-sectional studies***

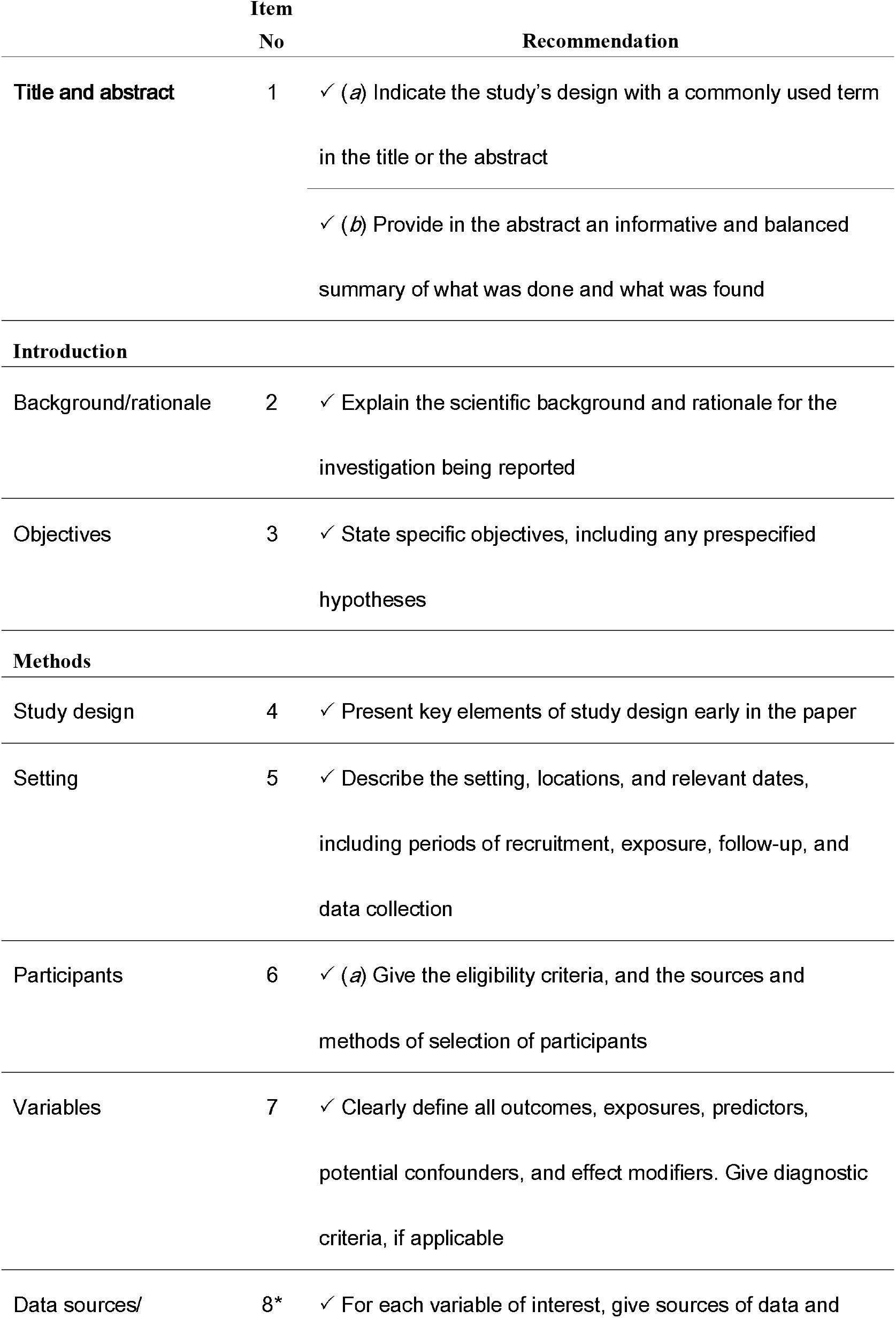
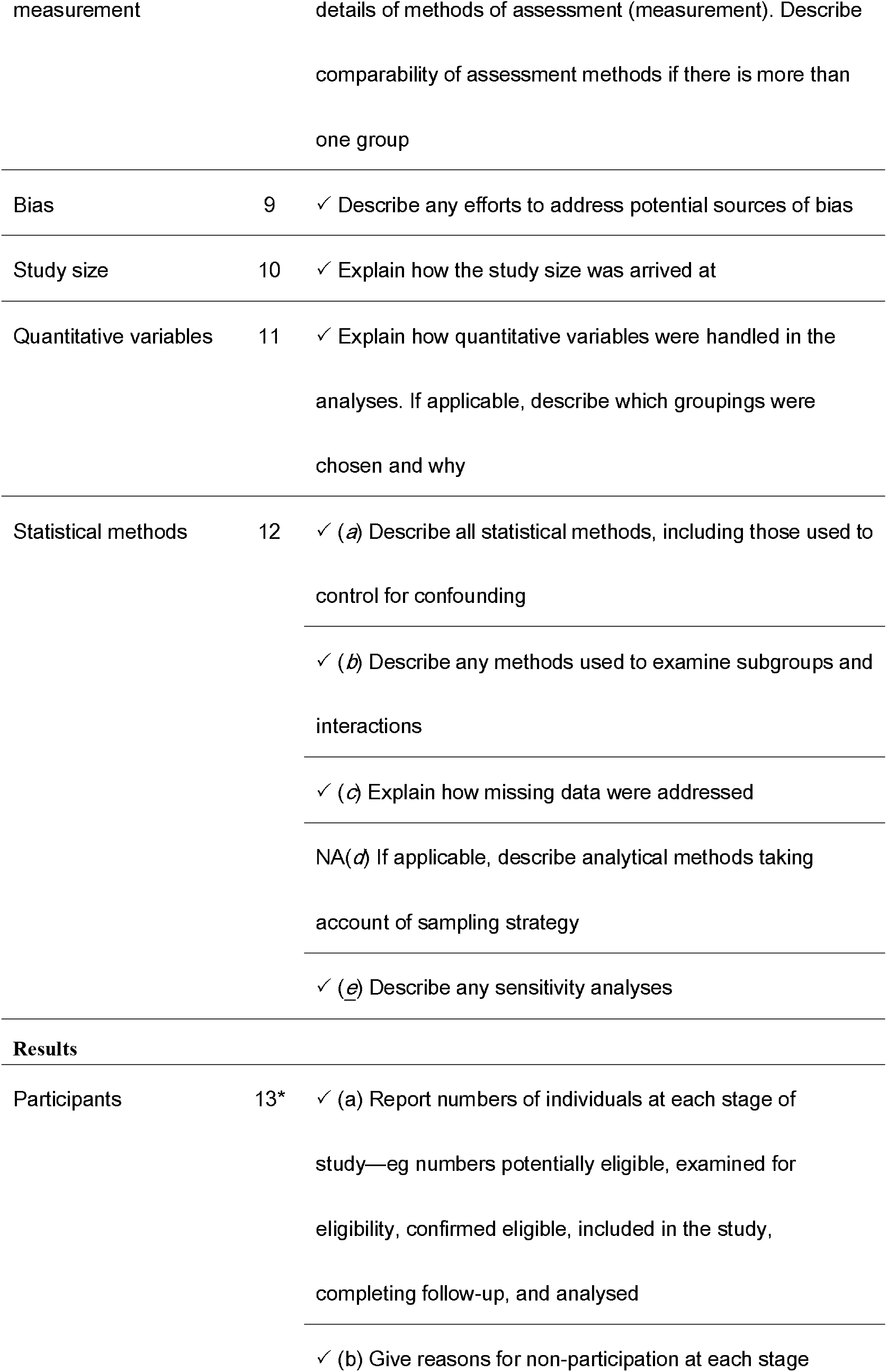
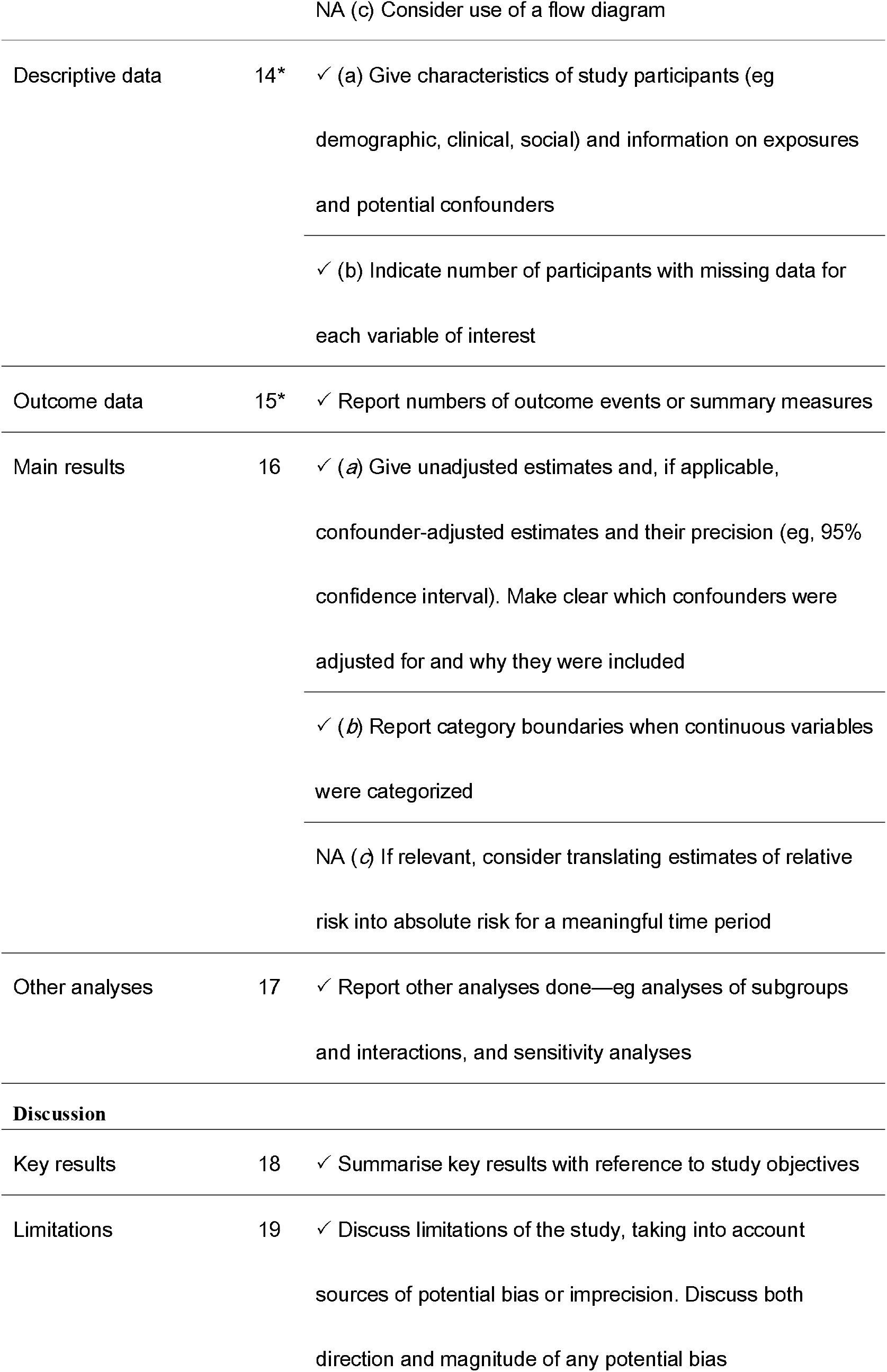
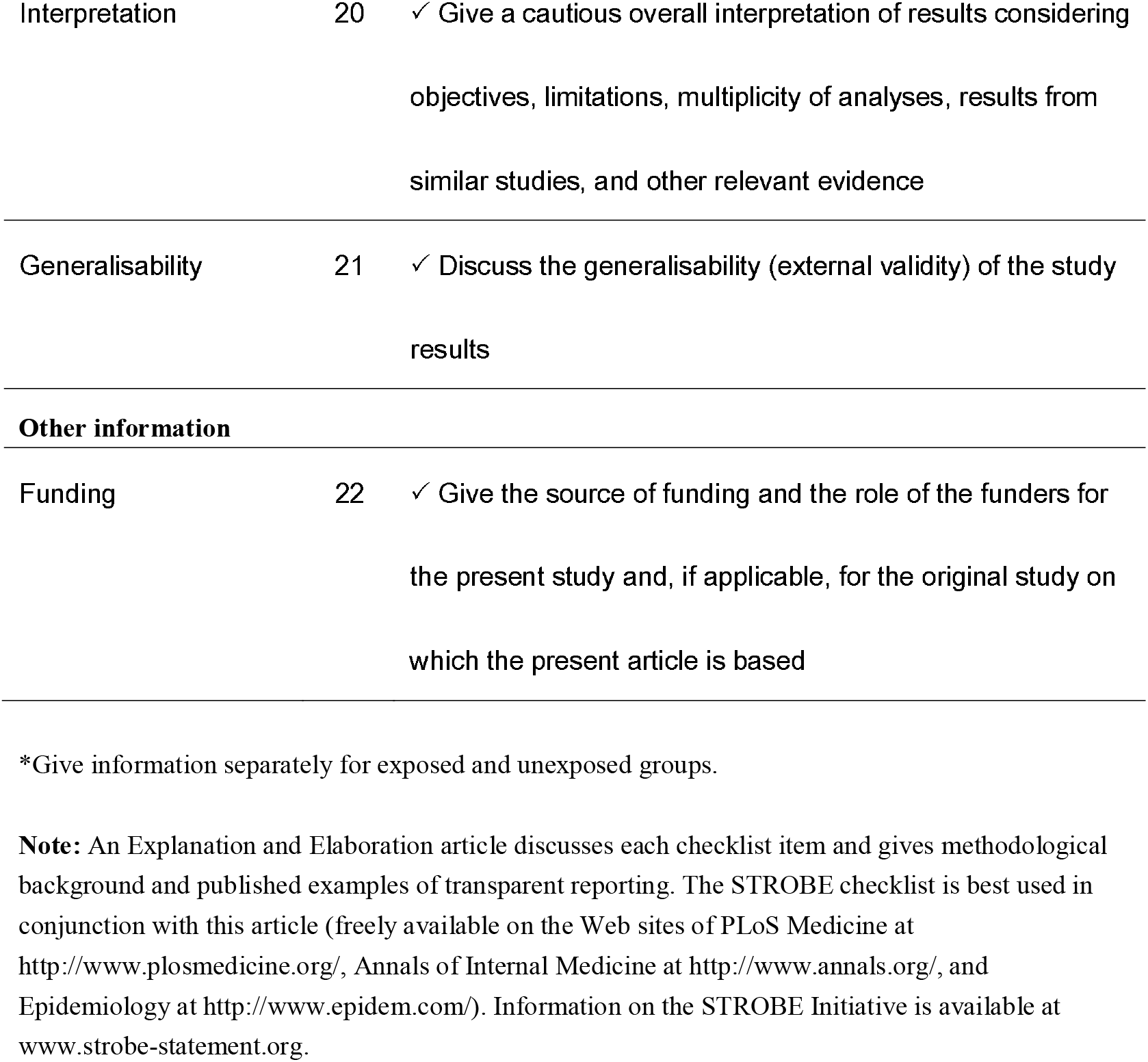

## Reference

1. Black CJ, Drossman DA, Talley NJ, Ruddy J, Ford AC. Functional gastrointestinal disorders: advances in understanding and management. The Lancet. 2020;396(10263):1664–74.

2. Sperber AD, Bangdiwala SI, Drossman DA, Ghoshal UC, Simren M, Tack J, et al. Worldwide Prevalence and Burden of Functional Gastrointestinal Disorders, Results of Rome Foundation Global Study. Gastroenterology. 2021;160(1):99–114.e3.

3. Drossman DA. Functional gastrointestinal disorders: history, pathophysiology, clinical features, and Rome IV. Gastroenterology. 2016;150(6):1262–79. e2.

4. Lin LD, Chang L. Using the Rome IV Criteria to Help Manage the Complex IBS Patient. Official journal of the American College of Gastroenterology | ACG. 2018;113(4):453–6.

5. Nakov R, Dimitrova-Yurukova D, Snegarova V, Uzunova M, Lyutakov I, Ivanova M, et al. Prevalence of Irritable Bowel Syndrome, Functional Dyspepsia and their Overlap in Bulgaria: a Population-Based Study. J Gastrointestin Liver Dis. 2020;29(3):329–38.

6. Park H. Functional gastrointestinal disorders and overlap syndrome in Korea. J Gastroenterol Hepatol. 2011;26 Suppl 3:12–4.

7. von Wulffen M, Talley NJ, Hammer J, McMaster J, Rich G, Shah A, et al. Overlap of Irritable Bowel Syndrome and Functional Dyspepsia in the Clinical Setting: Prevalence and Risk Factors. Dig Dis Sci. 2019;64(2):480–6.

8. Aziz I, Palsson OS, Törnblom H, Sperber AD, Whitehead WE, Simrén M. Epidemiology, clinical characteristics, and associations for symptom-based Rome IV functional dyspepsia in adults in the USA, Canada, and the UK: a cross-sectional population-based study. The lancet Gastroenterology & hepatology. 2018;3(4):252–62.

9. Carbone F, Fikree A, Aziz Q, Tack J. Joint Hypermobility Syndrome in Patients With Functional Dyspepsia. Clin Transl Gastroenterol. 2020;11(11):e00220.

10. Tanaka Y, Kanazawa M, Palsson OS, Tilburg MAV, Gangarosa LM, Fukudo S, et al. Increased Postprandial Colonic Motility and Autonomic Nervous System Activity in Patients With Irritable Bowel Syndrome: A Prospective Study. J Neurogastroenterol Motil. 2018;24(1):87–95.

11. Simrén M, Törnblom H, Palsson OS, van Tilburg MAL, Van Oudenhove L, Tack J, et al. Visceral hypersensitivity is associated with GI symptom severity in functional GI disorders: consistent findings from five different patient cohorts. Gut. 2018;67(2):255–62.

12. Burns G, Carroll G, Mathe A, Horvat J, Foster P, Walker MM, et al. Evidence for Local and Systemic Immune Activation in Functional Dyspepsia and the Irritable Bowel Syndrome: A Systematic Review. Am J Gastroenterol. 2019;114(3):429–36.

13. Drossman DA, Hasler WL. Rome IV-Functional GI Disorders: Disorders of Gut-Brain Interaction. Gastroenterology. 2016;150(6):1257–61.

14. Levy RL, Olden KW, Naliboff BD, Bradley LA, Francisconi C, Drossman DA, et al. Psychosocial aspects of the functional gastrointestinal disorders. Gastroenterology. 2006;130(5):1447–58.

15. Dipesh HV, Peter AP, Christopher JB, Lesley AH, Hazel AE, Maura C, et al. British Society of Gastroenterology guidelines on the management of irritable bowel syndrome. Gut. 2021;70(7):1214.

16. Gierk B, Kohlmann S, Kroenke K, Spangenberg L, Zenger M, Brähler E, et al. The somatic symptom scale–8 (SSS-8): a brief measure of somatic symptom burden. JAMA internal medicine. 2014;174(3):399–407.

17. Kanazawa M, Nakajima S, Oshima T, Whitehead WE, Sperber AD, Palsson OS, et al. Validity and reliability of the Japanese version of the Rome III diagnostic questionnaire for irritable bowel syndrome and functional dyspepsia. J Neurogastroenterol Motil. 2015;21(4):537.

18. Bouchoucha M, Devroede G, Fysekidis M, Rompteaux P, Sabate JM, Benamouzig R. Data Mining Approach for the Characterization of Functional Bowel Disorders According to Symptom Intensity Provides a Small Number of Homogenous Groups. Digestive Diseases. 2020;38(4):310–9.

19. Berens S, Engel F, Gauss A, Tesarz J, Herzog W, Niesler B, et al. Patients with Multiple Functional Gastrointestinal Disorders (FGIDs) Show Increased Illness Severity: A Cross-Sectional Study in a Tertiary Care FGID Specialty Clinic. Gastroenterol Res Pract. 2020;2020:9086340.

20. Polster AV, Palsson OS, Törnblom H, Öhman L, Sperber AD, Whitehead WE, et al. Subgroups of IBS patients are characterized by specific, reproducible profiles of GI and non-GI symptoms and report differences in healthcare utilization: A population-based study. Neurogastroenterology & Motility. 2019;31(1):e13483.

21. Polster A, Van Oudenhove L, Jones M, Öhman L, Törnblom H, Simrén M. Mixture model analysis identifies irritable bowel syndrome subgroups characterised by specific profiles of gastrointestinal, extraintestinal somatic and psychological symptoms. Alimentary Pharmacology & Therapeutics. 2017;46(5):529–39.

22. Siah KTH, Gong X, Yang XJ, Whitehead WE, Chen M, Hou X, et al. Rome Foundation-Asian working team report: Asian functional gastrointestinal disorder symptom clusters. Gut. 2018;67(6):1071–7.

23. Chey WD, Lembo AJ, Lavins BJ, Shiff SJ, Kurtz CB, Currie MG, et al. Linaclotide for Irritable Bowel Syndrome With Constipation: A 26-Week, Randomized, Double-blind, Placebo-Controlled Trial to Evaluate Efficacy and Safety. Official journal of the American College of Gastroenterology | ACG. 2012;107(11):1702–12.

24. Drossman DA, Chey WD, Johanson JF, Fass R, Scott C, Panas R, et al. Clinical trial: lubiprostone in patients with constipation-associated irritable bowel syndrome--results of two randomized, placebo-controlled studies. Aliment Pharmacol Ther. 2009;29(3):329–41.

25. Drossman DA. Functional Gastrointestinal Disorders: History, Pathophysiology, Clinical Features, and Rome IV. Gastroenterology. 2016;150(6):1262–79.e2.

26. Ghoshal UC. Marshall and Warren Lecture 2019: a paradigm shift in pathophysiological basis of irritable bowel syndrome and its implication on treatment. Journal of Gastroenterology and Hepatology. 2020;35(5):712–21.

27. Huang Z, Lyu Z, Hou Z, Wu Y, Huang J, Liu F, et al. Quantifying Liver-Stomach Disharmony Pattern of Functional Dyspepsia Using Multidimensional Analysis Methods. Evidence-Based Complementary and Alternative Medicine. 2020;2020.

28. Dowling NM, Bolt DM, Deng S, Li C. Measurement and control of bias in patient reported outcomes using multidimensional item response theory. BMC Medical Research Methodology. 2016;16(1):63.

29. Goldenberg JZ, Steel A, Day A, Yap C, Bradley R, Cooley K. Naturopathic approaches to irritable bowel syndrome: protocol for a prospective observational study in academic teaching clinics. Integrative Medicine Research. 2018;7(3):279–86.

30. Chalmers RP. Generating adaptive and non-adaptive test interfaces for multidimensional item response theory applications. Journal of Statistical Software. 2016;71(5):1–39.

31. Miwa H, Ghoshal UC, Fock KM, Gonlachanvit S, Gwee KA, Ang TL, et al. Asian consensus report on functional dyspepsia. Journal of gastroenterology and hepatology. 2012;27(4):626–41.

32. Gwee KA, Bak YT, Ghoshal UC, Gonlachanvit S, Lee OY, Fock KM, et al. Asian consensus on irritable bowel syndrome. Journal of gastroenterology and hepatology. 2010;25(7):1189–205.

33. Hu P. Consensus on diagnosis and treatment of irritable bowel syndrome (2007, Changsha). Chinese Journal of General Practitioners. 2008;7(5):298–300.

34. Li J, Chen J, Li Y. Consensus on the Diagnosis and Treatment of Functional Dyspepsia combined Chinese and Western Medicine (2017). Chinese Journal of Integrated Chinese and Western Medicine digestion. 2017(12):889–94.

35. Li J, Chen J, Tang X, Bian L. Consensus on the Diagnosis and Treatment of Irritable bowel Syndrome by Integrated Traditional Chinese and Western Medicine (2017). Chinese Journal of Integrated Chinese and Western Medicine digestion. 2018(3):227–32.

36. Francis CY, Morris J, Whorwell PJ. The irritable bowel severity scoring system: a simple method of monitoring irritable bowel syndrome and its progress. Alimentary pharmacology & therapeutics. 1997;11(2):395–402.

37. Li L, Wang H, Shen Y. Chinese SF-36 Health Survey: translation, cultural adaptation, validation, and normalisation. Journal of Epidemiology & Community Health. 2003;57(4):259–63.

38. Chalmers RP. mirt: A Multidimensional Item Response Theory Package for the R Environment. Journal of Statistical Software. 2012;48(6):1–29.

39. Chalmers RP. mirtCAT: computerized adaptive testing with multidimensional item response theory https://cran.r-project.org/web/packages/mirtCAT/index.html2017 [2020-3-8].

40. Rasmussen C. The infinite Gaussian mixture model. Advances in neural information processing systems. 1999;12:554–60.

41. Pohl D, Van Oudenhove L, Törnblom H, Le Nevé B, Tack J, Simrén M. Functional Dyspepsia and Severity of Psychologic Symptoms Associate With Postprandial Symptoms in Patients With Irritable Bowel Syndrome. Clin Gastroenterol Hepatol. 2018;16(11):1745–53 e1.

42. Koloski NA, Jones M, Talley NJ. Evidence that independent gut-to-brain and brain-to-gut pathways operate in the irritable bowel syndrome and functional dyspepsia: a 1-year population-based prospective study. Aliment Pharmacol Ther. 2016;44(6):592–600.

43. Larsson MB, Tillisch K, Craig AD, Engström M, Labus J, Naliboff B, et al. Brain responses to visceral stimuli reflect visceral sensitivity thresholds in patients with irritable bowel syndrome. Gastroenterology. 2012;142(3):463–72 e3.

44. Van Oudenhove L, Vandenberghe J, Dupont P, Geeraerts B, Vos R, Dirix S, et al. Regional brain activity in functional dyspepsia: a H(2)(15)O-PET study on the role of gastric sensitivity and abuse history. Gastroenterology. 2010;139(1):36–47.

45. McCullough RW. IBS, NERD and functional dyspepsia are immuno-neuronal disorders of mucosal cytokine imbalances clinically reversible with high potency sucralfate. Med Hypotheses. 2013;80(3):230–3.

46. Lackner JM. The Role of Psychosocial Factors in Functional Gastrointestinal Disorders. Frontiers of Gastrointestinal Research. 2014;33:104–16.

47. Byale A, Lennon RJ, Byale S, Breen-Lyles M, Edwinson AL, Gupta R, et al. High-Dimensional Clustering of 4000 Irritable Bowel Syndrome Patients Reveals Seven Distinct Disease Subsets. Clin Gastroenterol Hepatol. 2022.

48. Black CJ, Houghton LA, Ford AC. Latent class analysis does not support the existence of Rome IV functional bowel disorders as discrete entities. Neurogastroenterol Motil. 2022;34(11):e14391.

49. Holtmann GJ, Talley NJ. Inconsistent symptom clusters for functional gastrointestinal disorders in Asia: is Rome burning? Gut. 2018;67(11):1911–5.

50. Zinsmeister AR, Herrick LM, Saito Loftus YA, Schleck CD, Talley NJ. Identification and validation of functional gastrointestinal disorder subtypes using latent class analysis: a population-based study. Scandinavian Journal of Gastroenterology. 2018;53(5):549–58.

